# A mathematical model to investigate the transmission of COVID-19 in the Kingdom of Saudi Arabia

**DOI:** 10.1101/2020.05.02.20088617

**Authors:** Fehaid Salem Alshammari

## Abstract

Since the first confirmed case of SARS-CoV-2 coronavirus (COVID-19) in the 2*^nd^* day of March, Saudi Arabia has not report a quite rapid COVD-19 spread compared to America and many European countries. Possible causes include the spread of asymptomatic cases. To characterize the transmission of COVID-19 in Saudi Arabia, this paper applies a susceptible, exposed, symptomatic, asymptomatic, hospitalized, and recovered dynamical model, along with the official COVID-19 reported data by the Ministry of Health in Saudi Arabia. The basic reproduction number *R*_0_ is estimated to range from 2.87 to 4.9.

## 1 Introduction

As of April 22, 2020, more than 12772 cases and 114 deaths of coronavirus disease 2019 (COVID-19) caused by the SARS-CoV-2 virus had been confirmed in Saudi Arabia. Since the 4*^th^* of March [17], control measures have been implemented within Saudi Arabia to try to control the spread of the disease. Isolation of confirmed cases and contact tracing are crucial part of these measures, which are common interventions for controlling infectious disease outbreaks [26–28]. For example, the severe acute respiratory syndrome (SARS) outbreak SARS and Middle East respiratory syndrome (MERS), were controlled through tracing suspected cases and isolating confirmed cases because the majority of transmission occurred concurrent or after symptom onset [27–29].

However, it is unknown if transmission of COVID-19 can occur before symptom onset, which could decrease the effectiveness of isolation and contact tracing [26, 27, 29].

In this paper, the impact of asymptomatic COVID-19 cases on the spread of the disease will be considered using a modified version of the susceptible-exposed-infected-recovered (SEIR) dynamical model, along with the official COVID-19 data reported by the ministry of health in Saudi Arabia. Other main objectives of this paper include: estimating the basic reproduction number *(R*_0_) of COVID-19 in Saudi Arabia and how interacting with infected individual (symptomatic and asymptomatic) affect the estimated number, estimating the maximum required number of hospital beds and intensive care units (ICU).

## 2 Model establishment

The population will be divided into six categories: susceptible (*S*), exposed (*E*), symptomatic (*Y*), asymptomatic (*N*), hospitalized (*H*), and recovered (*R*) individuals (SEYNHR). Individuals moves from the susceptible compartment *S* to the exposed compartment *E* after interacting with infected individuals with transmission rates *β*_1_, *β*_2_, and *β*_3_ as shown in Figure 1. COVID-19 is known to have an incubation period, from 2 to 14 days, between exposure and development of symptoms [6,30]. After this period, exposed individual transits from the compartment E to either compartment *Y* at a rate *α*, or to compartment *N* at a rate *α*(1 − *γ*). An individual could move from compartment *N* to *Y* at a rate *K* if they show symptoms. Once an individual becomes infected with the coronavirus that causes COVID-19, that individual develops immunity against the virus with a rate Φ*_Y_* or the individual will be hospitalized with a rate of ϵ or die because of the disease with a rate of *μ*_1_. When individual becomes hospitalized, that individual receives treatment and develops immunity against the virus with a rate r or die because of the disease with a rate of *μ*_2_.

**Figure 1:**
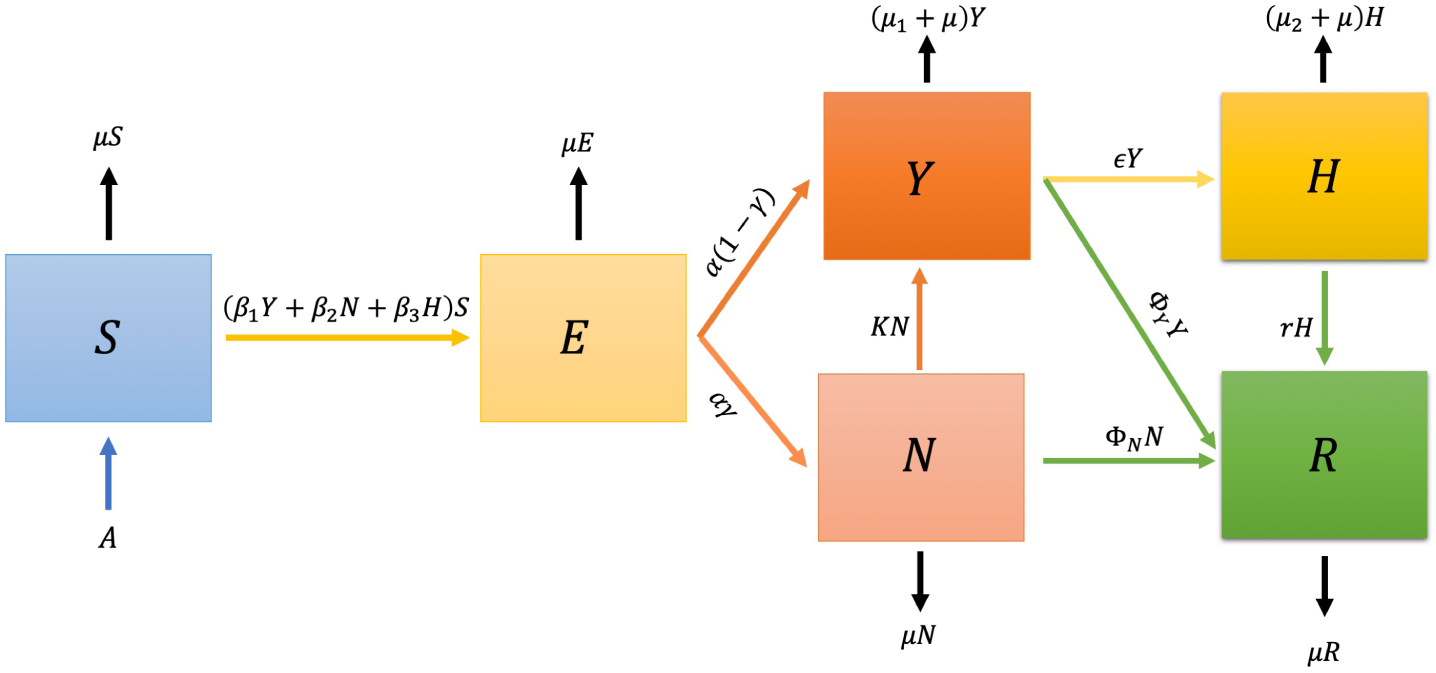
Schematic diagram of SEIHR compartment model. The arrows, except the black ones, represent progression from one compartment to the next.

As shown in Figure 1, the SEYNHR model has six compartments, and there fore a discrete dynamical system consisting six non-linear differential equations will be formed as the following:

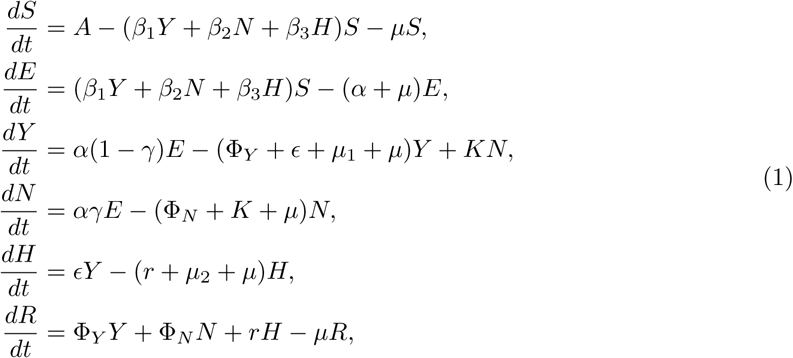

where *N*(*t*) = *S*(*t*) + *E*(*t*) + *Y*(*t*) + *N*(*t*) + *H*(*t*) + *R*(*t*). The next-generation matrix will be used to derive an analytical expression for the basic reproduction number (*R*_0_), for the compartmental model above. Calculating *R*_0_ is a useful metric for assessing the transmission potential of an emerging COVID19 in Saudi Arabia.

## 3 Basic reproduction number *R*_0_

An important concept in epidemiology is the basic reproduction number, defined as “the expected number of secondary cases produced, in a completely susceptible population, by a typical infective individual” [10]. The next generation method will be used to calculate *R*_0_ [11]. The system in Equation 1 can be rewritten as follows

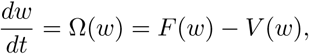

where F: = (*F*1, *F*2, *F*3, *F*4, *F*5, *F*6)*^T^* and *V*: = (*V*1, *V*2, *V*3, *V*4, *V*5, *V*6)*^T^*, or more explicitly

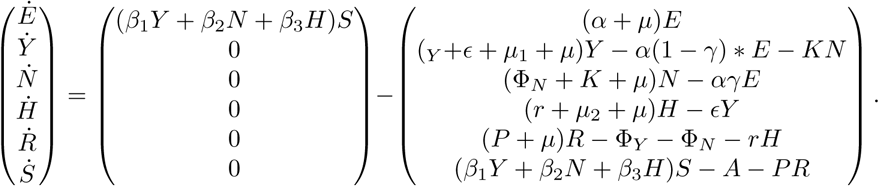

The Jacobian matrices of *F* and *V* evaluated at the disease-free equilibrium (DFE) of the system in Equation 1, 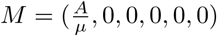, are given by

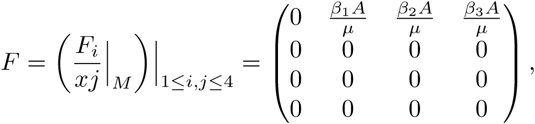

and

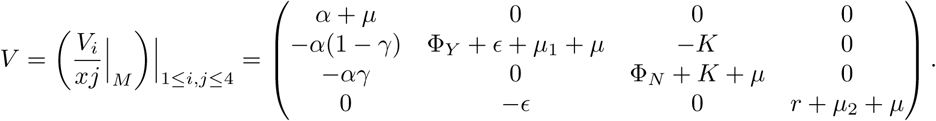

Direct calculations show that

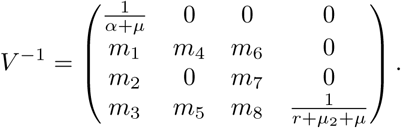

where

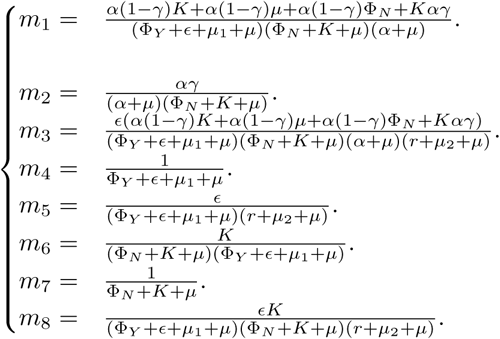

Denoting the 4×4 identity matrix by 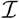, the characteristic polynomial Γ(λ) of the matrix *FV*^−1^ is given by

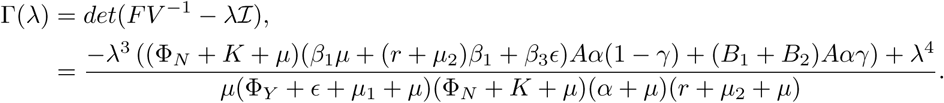

where

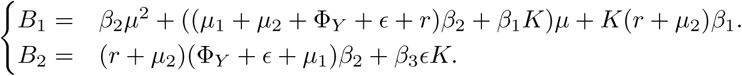

The solutions λ_1,2,3,4_ are given by

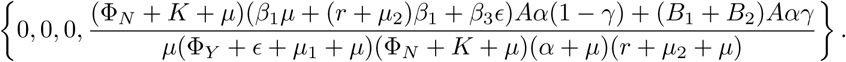

Therefore, the reproduction number for the *SEYNHR* model in Equation 1 is given by

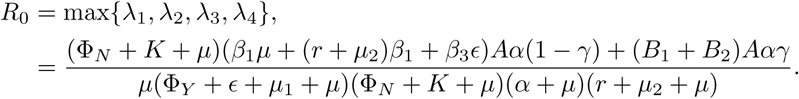

## 4 Results

At the initial time *t* = 0, if we set the initial population *N*(0) = 1, then *N*(0) = *S*(0)+*E*(0)+*Y*(0)+*N*(0)+*H*(0)+*R*(0) = 1. The transition probabilities between states are all in the range of [0, 1], i.e.,

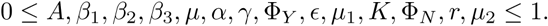

I fitted the model in Equation 1 to the published data from the Ministry of Health in Saudi Arabia (MOH) from 2^nd^ day of March until the 14^th^ of April to estimate values for the unknown parameters *β*_1_, *β*_2_, *β*_3_ and *K* using MATLAB. The results are shown in the upper panel of Figure 2. Increasing the value of *K* from 0.009/day requires increasing the value of *β*_1_, *β*_2_ and *β*_3_ from 0.267, 0.53 and 0.13 (day x individual)^−1^ to 0.5, 1 and 0.26 (day x individual)^−1^, respectively. All other parameters and their descriptions are given in Table 1. I assumed that the mean asymptomatic infectious period is the same as the mean symptomatic infectious period because there is no estimation available in the literature [9, 29]. Based on those estimated, assumed and measured values, the basic reproduction number *R*_0_ is estimated to range from 2.87 to 4.9. The variation of the basic reproduction number *R*_0_ for different values of *β*_1_, *β*_2_, *Φ_Y_, Φ*_N_ and *K* are shown in the heat maps in Figure 3. The upper heat map of Figure 3 shows that practicing physical distancing could significantly reduce the value of *R*_0_ and hence control the spread of the disease.

**Figure 2:**
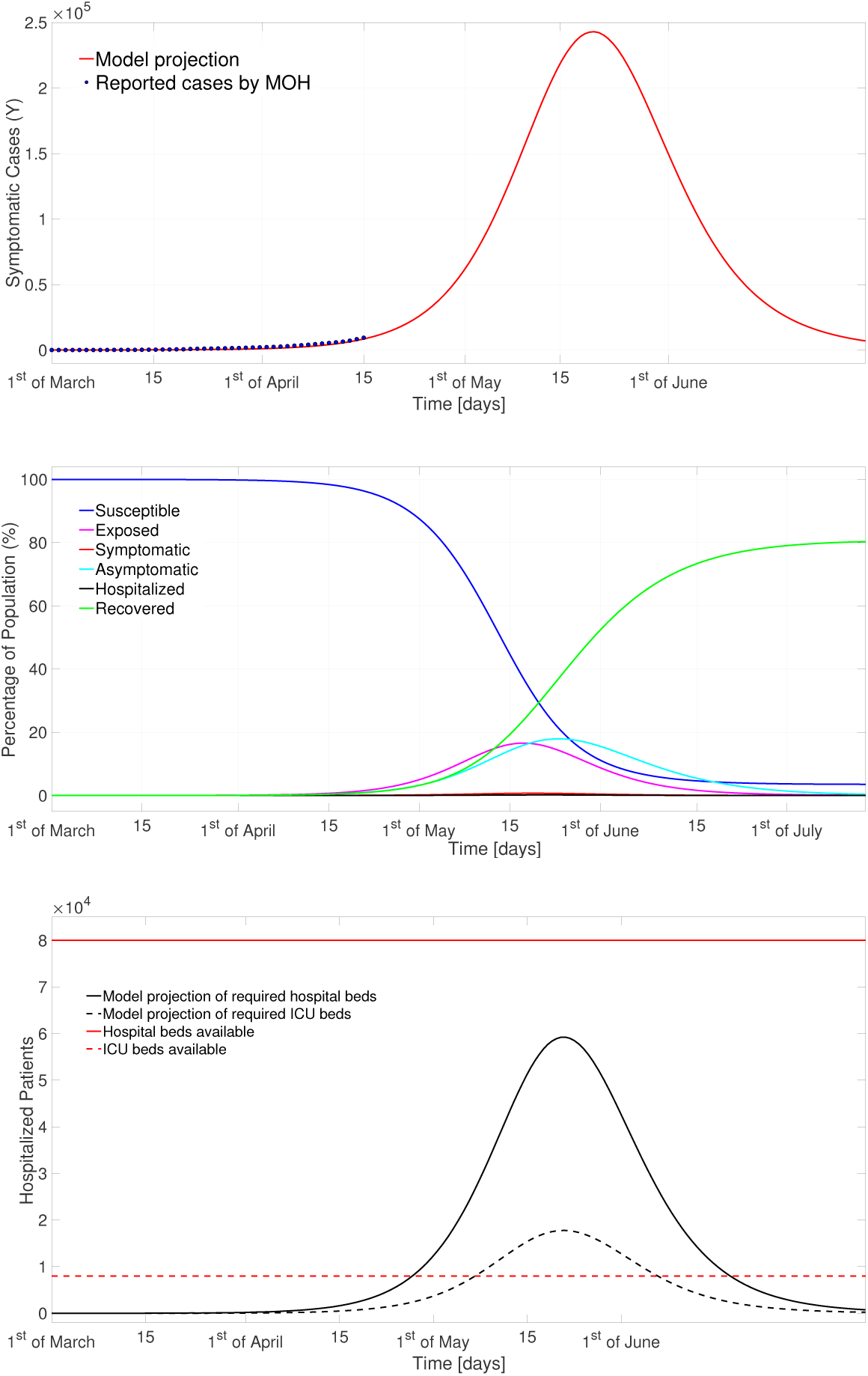
Numerical results of the SEYNHR model with the parameter listed on Table 1, where *β*_1_ = 0.35, *β*_2_ = 0.7, *β*_3_ = 0.18 and *K* = 0.009. The upper figure shows the estimated number of symptomatic COVID-19 cases, with the published data by the Ministry of Health in Saudi Arabia of confirmed COVID-19 cases (blue circles). The center figure shows the estimated susceptible, exposed, symptomatic, asymptomatic, hospitalized, and recovered sub-populations. The lower figure shows estimations of the hospitalized cases and the required ICU beds (black dashed line). The red line represents the number of hospital beds available, while the red dashed line represents the number of ICU beds available in Saudi Arabia.

**Figure 3:**
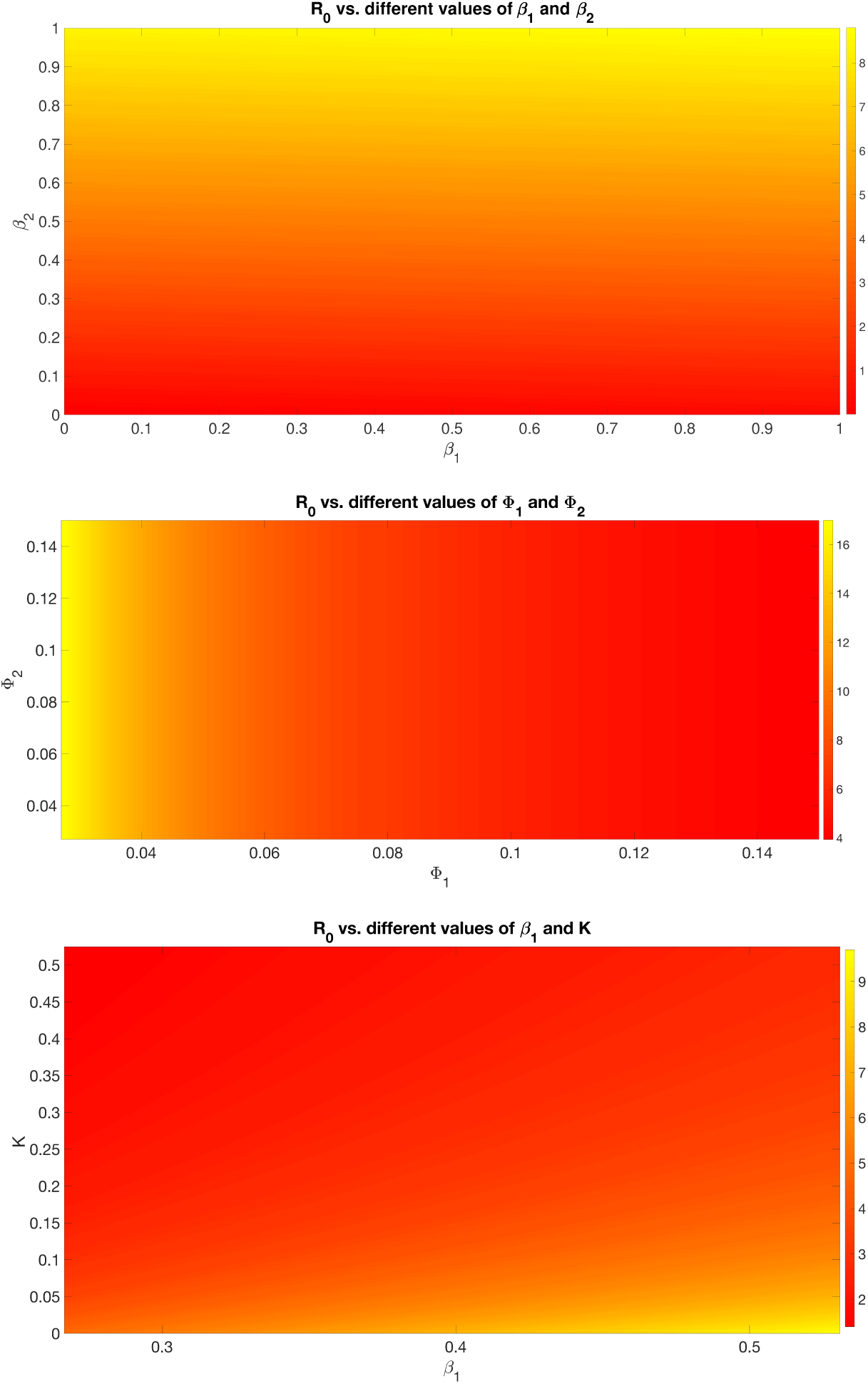
Heat maps showing the variation of *R*_0_ for different parameter values: the upper heat map shows the variation of *R*_0_ for different values for *β*_1_ and *β*_2_, in the center the heat map shows the variation of *R*_0_ for different values for and Φ*_N_* and the lower heat map shows the variation of *R*_0_ for different values for *β*_1_ and *K*.

**Table 1:** Parameters used in the simulations.

| Parameter | Description | Dimension | Value | Source |
| --- | --- | --- | --- | --- |
| $N$ | Population of Saudi Arabia in 2019. | Individual | 34218169 | [12] |
| $A$ | Birth rate in Saudi Arabia in 2019. | Individual/day | 1650 | [12] |
| $\mu$ | Death rate in Saudi Arabia in 2019. | day<sup>-1</sup> | $3.7 \times 10^{-5}$ | [12] |
| $\beta_1$ | Effective contact rate from symptomatic to susceptible. | day<sup>-1</sup> | [0.267, 0.5] | fitted |
| $\beta_2$ | Effective contact rate from asymptomatic to susceptible. | day<sup>-1</sup> | [0.53, 1] | fitted |
| $\beta_3$ | Effective contact rate from hospitalized to susceptible. | day<sup>-1</sup> | [0.13, 0.26] | fitted |
| $\alpha^{-1}$ | Mean latent period. | days | 5.1 | [13] |
| $\gamma$ | Probability of becoming asymptomatic upon infection. | n/a | 0.86 | [14] |
| $\Phi_Y^{-1}$ | Mean symptomatic infectious period. | days | 8 | [15] |
| $\epsilon$ | Rate of symptomatic becomes hospitalized. | day<sup>-1</sup> | 0.125 | [17] |
| $\mu_1$ | Death rate of symptomatic patients. | day<sup>-1</sup> | 0.392 | [16] |
| $K$ | Rate of asymptomatic becomes symptomatic. | day<sup>-1</sup> | [0.009, 0.125] | fitted |
| $\Phi_N^{-1}$ | Mean asymptomatic infectious period. | days | 8 | assumed |
| $r$ | Rate of recovered hospitalized patients. | day<sup>-1</sup> | 0.1 | [6] |
| $\mu_2$ | Death rate of hospitalized patients. | day<sup>-1</sup> | 0.392 | [17] |

The center panel of Figure 2 shows that about 18% of the entire Saudi population will be asymptomatic in the last week of May 2020 and about 17% will be exposed in the third week of May. The percentage of the entire population being symptomatic at anytime will not exceed 1%, which is estimated to occur in the third week of May. Moreover, about 60000 hospital beds and 18000 ICU beds are required (30% of the hospitalized cases [6]) immediately after the second week of May. Currently, the Ministry of Health designated 25 hospitals for COVID-19 infected patients with up to 80,000 beds and 8000 intensive care units (ICU) beds [25] and therefore extra 10000 ICU beds could be required.

## 5 Discussion

The parameter with the hight degree of uncertainty are the effective contact rates from symptomatic to susceptible *β*_1_, from asymptomatic to susceptible *β*_2_ and from hospitalized to susceptible *β*_3_ (expected to be a fraction of *β*_1_ because of the protective measures in hospitals), as well as the mean infectious periods for symptomatic 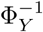 and asymptomatic 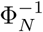 individuals. I estimated a maximum value of *β*_1_ to be 0.5 which is one half of the value reported by Li *et al*. [18]. This could be a reasonable estimation as we have not seen similar scenario in Saudi Arabia after 5 weeks since reaching 100 confirmed cases on the 14th of March (week 7 since the first case) as we have seen in many other countries like China, America and different European countries in the same timescale. This could be a result of the precautionary measures taken by the Saudi authorities, including closure of schools and universities that started as early as the 8*^th^* of March (six days after the first confirmed COVID-19 case in Saudi Arabia). Based on the above estimation for *R*_0_, the center panel of Figure 3 suggests that the infectious period for symptomatic patient could be in the range from 6.6 to 12.5 days (i.e., Φ*_Y_* ∊ [0.08, 0.15]) and the infectious period for asymptomatic patient could be in the range from 6.6 to 25 days (i.e., Φ*_N_* ∊ [0.04,0.15]). The infectious period for symptomatic cases are consistent with what is being observed clinically [30].

In reality, *R*_0_ is not a biological constant; it could fluctuate daily depending on environmental and social factors such as percentage of entire susceptible population wearing suitable medical mask and practicing physical distancing. In the literature, estimates of *R*_0_ vary greatly: from 1 to 6 [18–24] up to 26.5 [9]. This variation is because of different assumptions and factors they had considered in the calculations. In general, considering asymptomatic infection sub-population will increase the estimated values of *R*_0_.

## 6 Conclusion

The contribution of undocumented COVID-19 infections (asymptomatic cases) on the transmission of the disease deserves further studies and investigations. This paper shows that asymptomatic cases of COVID-19 will drive the growth of the pandemic in Saudi Arabia. Therefore, more testing is needed to identify COVID-19 patients (symptomatic and asymptomatic) and to contain the spread of the disease.

## Data Availability

The data used in my paper is the total number of confirmed cases of COVID-19 reported by the ministry of Health in Saudi Arabia from 2nd of March until the 21st of April 2020. Data supporting this paper are open and can be accessed through:
Ministry of Health COVID 19 Dashboard. Saudi Arabia; 2020 [https://covid19.moh.gov.sa].

https://covid19.moh.gov.sa

